# An investigation of the reach of the Interim Canada Dental Benefit for children under 12 years of age

**DOI:** 10.1101/2025.04.08.25325411

**Authors:** Robert J. Schroth, Vivianne Cruz de Jesus, Carol Youssef, Olubukola O. Olatosi, Victor HK Lee, Saif Goubran, Eefa Khan, Anil Menon

## Abstract

**Introduction:** The Interim Canada Dental Benefit (CDB), introduced in 2022, provided financial assistance to families with children < 12 years. This study analyzed data from the Canada Revenue Agency (CRA) during the program’s entirety.

**Methods:** Data were accessed from the CRA for applicants and covered both the first (October 1, 2022–June 30, 2023) and second (July 1, 2023–June 30, 2024) periods. Rates of participation were calculated using population data from Statistics Canada. Adjusted rates were calculated based on the proportion of children without private dental insurance, and without private or public insurance.

**Results:** Over the 21 months of the Interim CDB, 408,240 regular applications were made and $401M distributed to Canadian families. More applications were made during period 1 (P1) than period 2 (P2), but more funding distributed in P2; $197M for 204,270 applications in P1 and $203M for 203,970 applications in P2. Overall, 321,000 children received the Interim CDB in P1 and 328,040 in P2. Provinces with highest rates of child participation included Manitoba, Ontario, Nova Scotia, and Saskatchewan. The highest adjusted rates based on the proportion of children without private or public insurance were Nova Scotia (673.3/1000 P1 and 717.8/1000 P2), Northwest Territories (618.4/1000 P1 and 573.2/1000 P2), and Saskatchewan (495.1/1000 P1 and 528.3/1000 P2)

**Conclusions:** Regions with access to care challenges had higher rates uptake of the Interim CDB when adjusting for the lack of private or public insurance. Findings from this study may help inform policy decisions and reach of the CDCP.

## 1 Introduction

Oral health is a significant determinant of overall health, yet disparities in dental care access are prevalent across Canada. These disparities are particularly common among children from priority populations who often experience a higher caries prevalence.^1^ Historically, the Canadian healthcare system did not include dental care.^2, 3^ Until recently, oral health remained largely outside of Canada’s publicly funded healthcare system.^4^

The limited public funding in Canada for dental care targets specific groups.^4^ The federal government provides funding for dental services to specific population groups (e.g., registered First Nations and Inuit Peoples, Canadian Armed Forces).^5^ Provincial governments also fund dental care, particularly for low-income children, social welfare recipients, individuals with disabilities, and those with craniofacial disorders.^5^ Additionally, some municipalities often share costs for dental care for low-income children and social welfare recipients with provinces, and they independently provide care for low-income seniors.^5^

Despite these government investments, access to oral health care remains a challenge for many Canadians. Vulnerable groups, including low-income households, children, seniors, Indigenous populations, and individuals with disabilities, face barriers to dental care, such as financial constraints, transportation issues, and limited services in rural and remote regions.^6^ Statistics show that while three-quarters of Canadians visit a dental professional annually, many still lack consistent access, largely due to cost.^7^

The Interim Canada Dental Benefit (CDB), introduced in October 2022, was a critical first step in addressing these access barriers for children under 12 years from low-income families.^8, 9^ The Interim CDB was implemented as a precursor to the Canadian Dental Care Plan (CDCP), providing eligible families of children < 12 years of age with financial support towards their children’s dental expenses.

Families earning under $90,000 annually were eligible to receive the Interim CDB for children under 12 years. Families with private insurance or employer-sponsored plans were ineligible. Applicants needed to have filed income taxes for the previous year. However, applicants who received government dental benefits, such as the Non-Insured Health Benefits (for registered First Nations and Inuit Peoples) or Employment and Income Assistance (i.e., social assistance), were also eligible for the Interim CDB if their child’s dental expenses were not fully reimbursed. Based on family income, financial support ranged from $260 to $650 per eligible child, with payments managed by the Canada Revenue Agency (CRA).^8-10^ The Period 1 (P1) of the Interim CDB covered eligible dental treatment between October 1, 2022, and June 30, 2023, and Period 2 (P2) covered treatment from July 1, 2023, to June 30, 2024.

As the Interim CDB ended June 30, 2024, a new chapter in Canadian dental care began with applications for the CDCP on June 27, 2024. The CDCP, with its expanded coverage to a broader range of uninsured Canadians with family incomes of less than $90,000, represents a significant policy shift towards more inclusive universal dental coverage. This transition is a testament to the continuous improvement and evolution of dental care policy in Canada. The purpose of this study was to analyze aggregated data from the Government of Canada on applications made and accepted during the entire 21 months of the Interim CDB, to evaluate its reach to gain insights into dental care needs and barriers faced by low-income Canadian families.

## 2 Methods

The study used aggregated data from the CRA regarding approved applications, unique applicants, and total funds distributed from October 2022 to June 2024. Public data were accessed from the Government of Canada Open Data Portal: https://open.canada.ca/data/en/dataset/69035265-2714-4ffa-af3f-fa850209b616 and https://www150.statcan.gc.ca/t1/tbl1/en/cv.action?pid=1310092001. Ethics approval was not required as it involved exclusively de-identified aggregated data publicly accessible from the government of Canada. The hypothesis was that Interim CDB uptake would be highest among lower-income families and regions with lower rates of private or public dental insurance coverage.

Data for this study were accessed from the CRA for applicants as of June 30, 2024, and assessed as of July 26, 2024. Data covered the entire first period (October 1, 2022–June 30, 2023) and 12 months of the second period (July 1, 2023–June 30, 2024). Data were segmented into four periods: Period 1 (P1), Period 2 (P2), Additional Period 1 (AP1), and Additional Period 2 (AP2). P1 includes applicants who applied, received the Interim CDB and sought dental care for their child(ren) between October 1, 2022, and June 30, 2023 (16). The P2 data include applicants who had dental treatment, applied, received the benefit, and sought dental care for their child(ren) between July 1, 2023, and June 30, 2024. AP1 data include applicants whose children had dental treatment with costs that exceeded $650 between October 1, 2022 and June 30, 2023. These applicants were eligible to apply and receive an additional $650 within the second pay period but would not be eligible to apply for the P2 of the Interim CDB. AP2 data include applicants who had children whose cost for dental treatment exceeded $650 between July 1, 2023, and June 30, 2024. They qualified to apply and receive an additional $650 within the second period, granted they had not applied for and received the Interim CDB within P1.

The analysis considered provincial/territorial variations, rates of child participation, and family net income distributions. New data for this follow-up also included additional metrics, such as changes in child participation levels and financial trends over the program’s lifetime. Descriptive statistics were calculated using Microsoft Excel, with data visualizations highlighting changes across all population segments.

The Canada Dental Benefit file provided the number of applications and the applicant’s province/territory of residence. Each unique applicant represented an individual, potentially applying for multiple children. Residence was determined as of the application date. All amounts were rounded to the nearest thousand dollars and counts to the nearest ten.

Rates of children with the Interim CDB per 1000 were calculated by dividing the number of children with the benefit by the number of Canadians aged 0-11, by province or territory, based on census 2021 data available from Statistics Canada.^8, 9, 11^ Additionally, we calculated adjusted rates of children without private dental insurance who received the Interim CDB per 1000. Provincial data on the percentage of children 0-17 years with private dental insurance was obtained from the Canadian Oral Health Survey (COHS) 2023-2024: Statistics Canada Table 13-10-0920-01 Selected indicators of dental insurance coverage by age group and gender, released October 23, 2024. This permitted us to calculate the percentage of children 0-17 years in each province without private insurance, which was then used to calculate adjusted rates for the total population of children 0-11 years of age in each province to reflect those without private insurance. Additionally, we determined the percentage of children 0-17 years with private or public dental insurance in each province to calculate adjusted rates for uninsured children 0-11 years (i.e., no private or public insurance). Data on private dental and public insurance for children in the three territories came from the Canadian Health Survey on Children and Youth (CHSCY) 2019, which was used to calculate the adjusted rates among the uninsured. Maps were generated using MapChart^©^ (https://www.mapchart.net/canada.html).

## 3 Results

Over the 21 months of the Interim CDB, 452,460 applications were made, and $440.7M was distributed to families. More applications were made during P1 than P2, but more funding was distributed in P2; $197M for 204,270 applications in P1 and $203M for 203,970 applications in P2. Overall, 321,000 children received the Interim CDB in P1, and 328,040 children received the Interim CDB in P2.

Ontario, Quebec, Alberta, and British Columbia had the highest number of approved applications and participants (Table 1), reflecting their larger populations and potentially greater need for dental benefits. Ontario consistently accounted for the most approved applications across all periods, reflecting its population size. The total approved amount was $90.6M and $92.0M for P1 and P2, respectively. Quebec reported a decline in the number of applications between the periods, from 37,830 in P1 to 36,560 in P2, while the amount distributed decreased accordingly. Meanwhile, the fewest applications came from the territories: Yukon, Nunavut, and the Northwest Territories (Table 1).

Data on the number of applications and amount distributed during AP1 and AP2 appear in Table 1. Overall, there were 16,680 applications made for additional funding in P1 and 27,540 in P2, with 24,660 and 39,300 children approved, respectively. The total amount distributed through this additional funding was $15.1M in AP1 and $24.6M in AP2. The majority of additional applications were from residents of Ontario (46.9% in AP1 and 42.9% in AP2) followed by Alberta (9.8% in AP1 and 10.1% in AP2). Similarly, most additional funds dispersed were highest in Ontario (47.0% in AP1 and 43.1% in AP2), followed by Alberta (14.1% in AP1 and 14.3% in AP2). Likewise, of all the children approved for additional funding, most were from Ontario (46.8% in AP1 and 42.8% in AP2), followed by Alberta (14.2% in AP1 and 14.4% in AP2).

Table 2 reports the number of children receiving the Interim CDB and the rate of child participation by province and territory during P1, P2, AP1, and AP2. Provinces with the highest rates of child participation included Ontario, Manitoba, Nova Scotia, and Saskatchewan (Figure 1a). Interestingly, all demonstrated an increase in rates from P1 to P2: Ontario (82.5/1,000 in P1 and 83.2/1,000 in P2), Manitoba (77.1/1,000 in P1 and 84.3/1000 in P2), Nova Scotia (73.4/1,000 in P1 and 78.2/1000 in P2), and Saskatchewan (72.3/1,000 in P1 and 77.1/1000 in P2) (Table 2 and Figure 1a). In Nunavut, participation increased from 22.1/1,000 to 28.2/1,000.

**Figure 1a.**
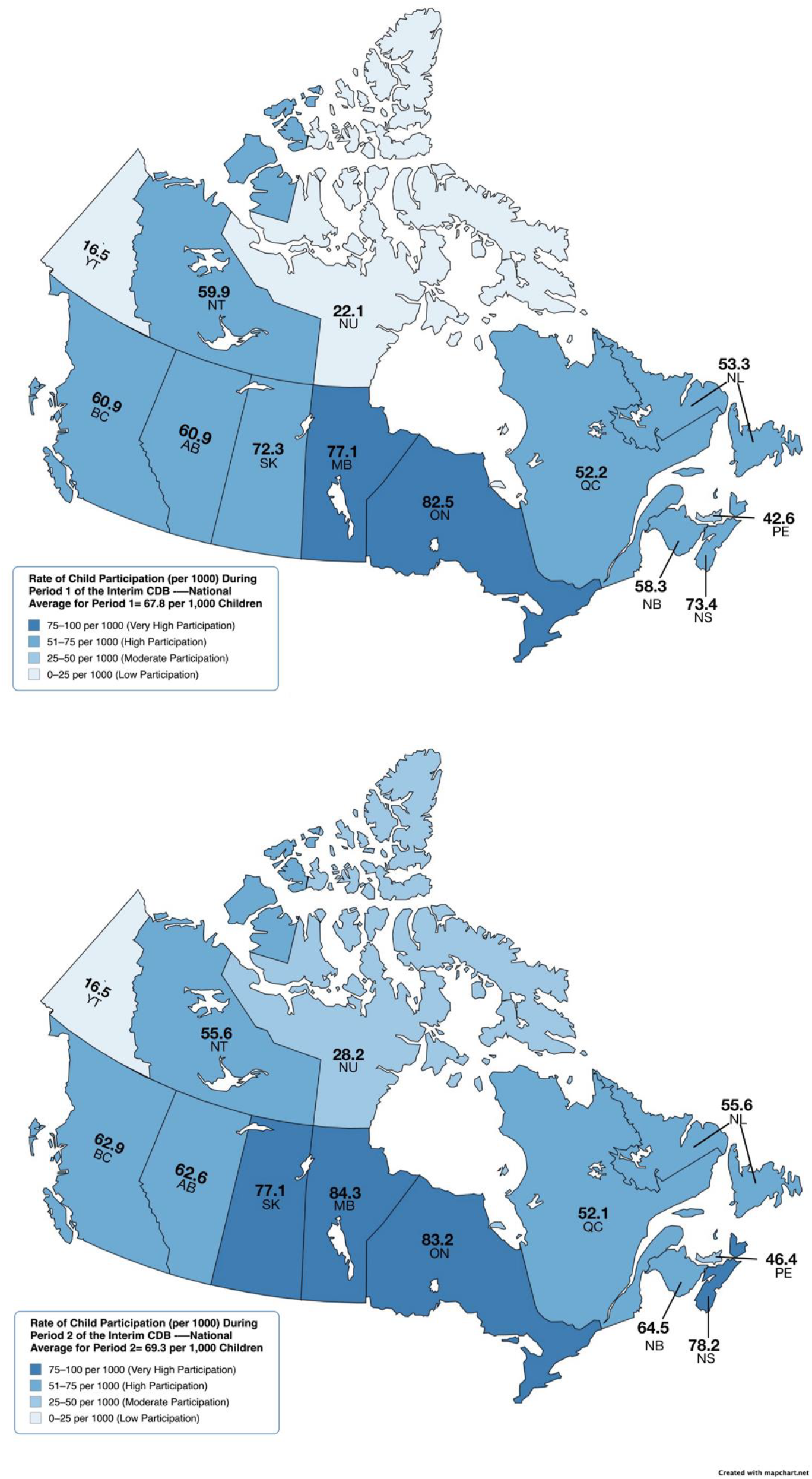
Rate of Child Participation (per 1000) During Period 1 and Period 2 of the Interim CDB

**Figure 1b.**
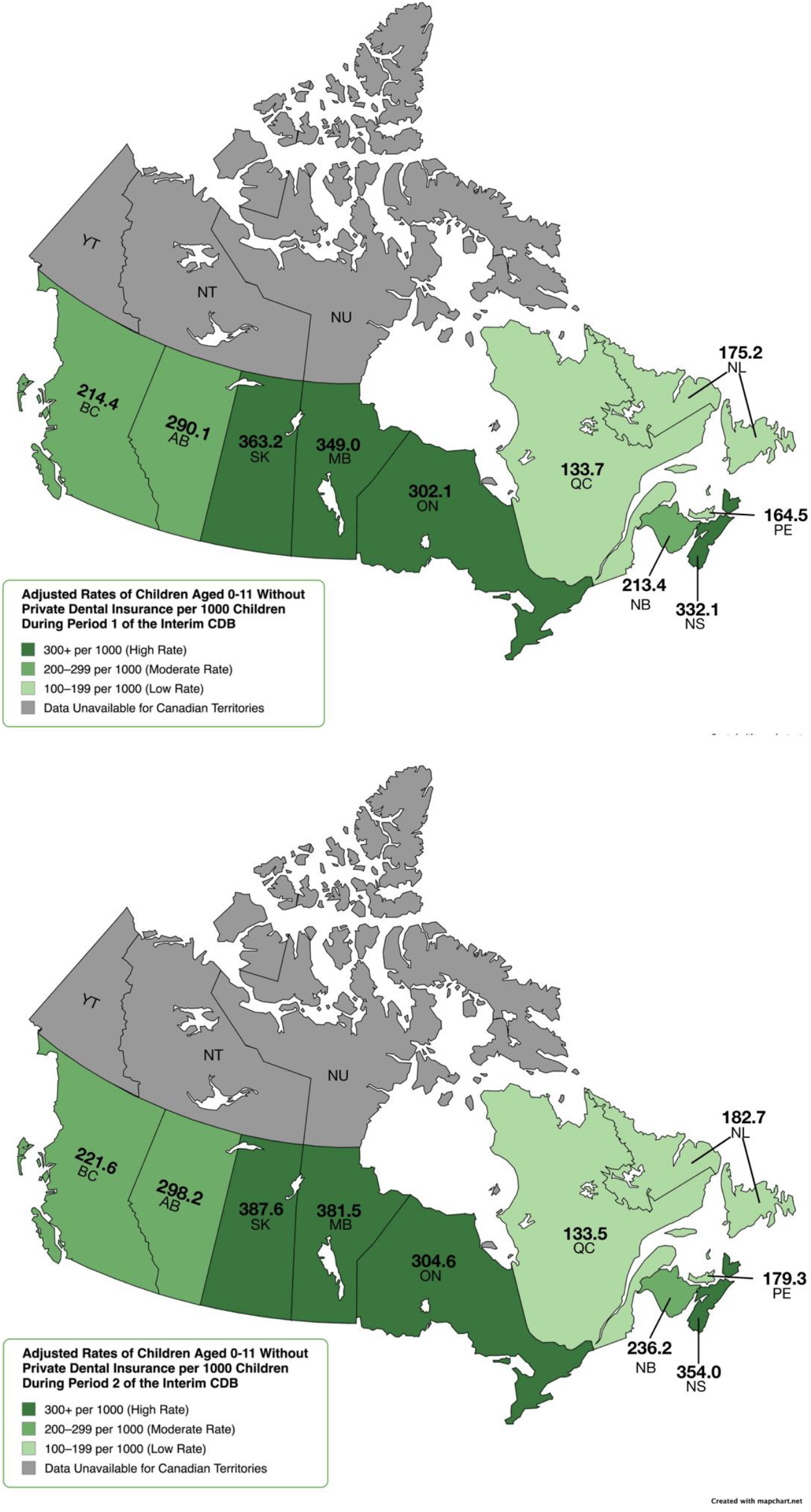
Adjusted rates of children aged 0-11 without private dental insurance per 1000 children during Period 1 and Period 2 of the Interim CDB

**Figure 1c.**
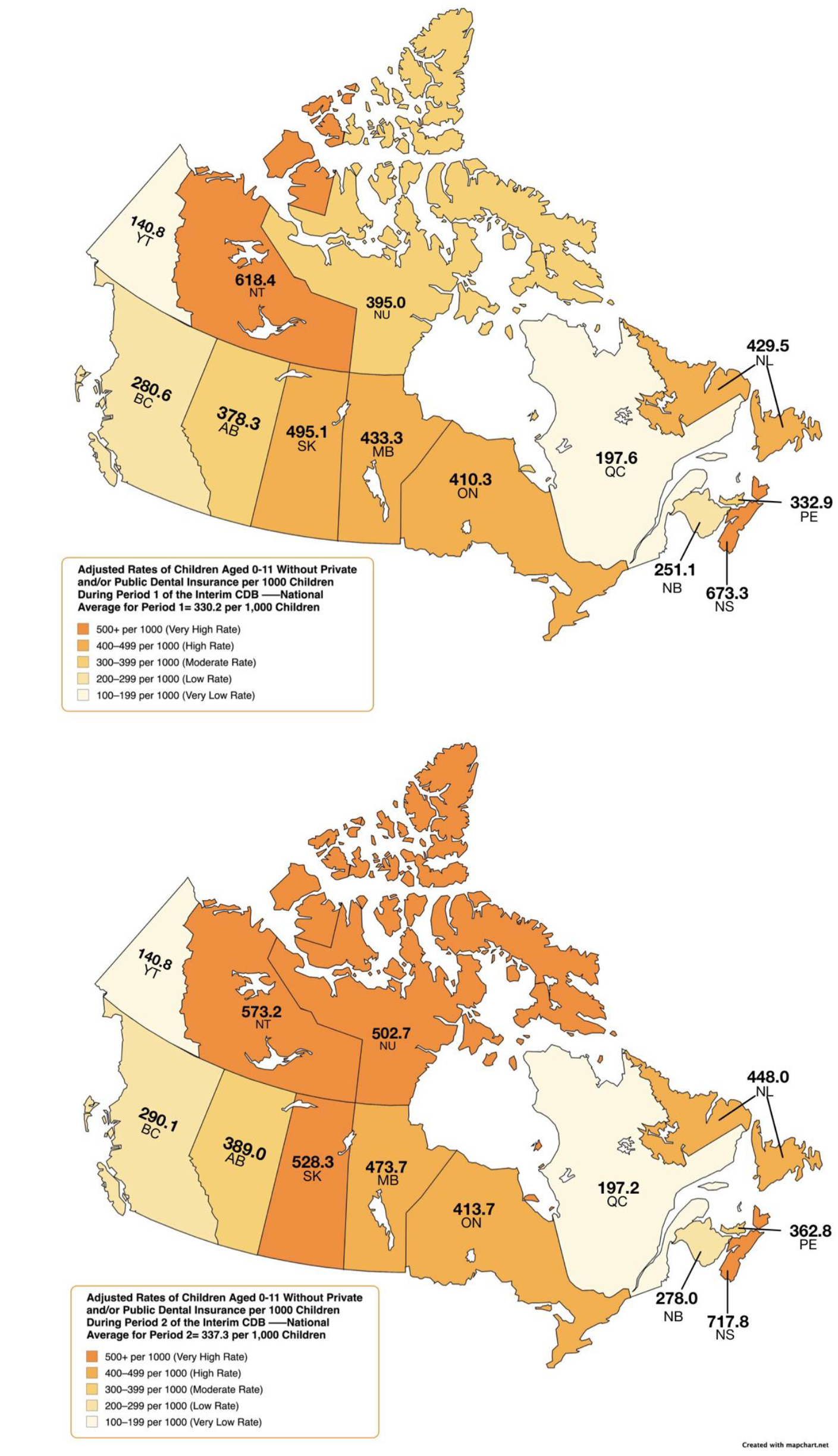
Adjusted rates of children aged 0-11 without private and/or public dental insurance per 1000 children during Period 1 and Period 2 of the Interim CDB

Adjusted rates of child participation limited to those without private dental insurance (obtained from the Canadian Oral Health Survey) revealed that Saskatchewan (363.2/1000) had the highest uptake in P1, followed by Manitoba (349.0/1000), Nova Scotia (332.1/1000), and Ontario (302.1/1000) (Figure 1b). Adjusted rates in P2 were slightly higher; Saskatchewan (387.6/1000), Manitoba (381.5/1000), Nova Scotia (354.0/1000), and Ontario (304.6/1000) (Figure 1b).

Adjusted rates of participation among those without private or public dental insurance (obtained from the Canadian Oral Health Survey for provinces and from the Canadian Health Survey on Children and Youth (CHSCY) 2019 for territories) revealed that the Canadian average rate of uptake was 330.2/1000 in P1 and 337.3/1000 in P2. Nova Scotia had the highest uptake rate in P1 (673.3/1000) followed by the Northwest Territories (618.4/1000), Saskatchewan (495.1/1000), Manitoba (433.3/1000), and Newfoundland and Labrador (429.5/1000) (Figure 1c). Seven provinces and territories saw an increase in the rate of uninsured children receiving the Interim CDB from P1 to P2, but rates in the Yukon and Quebec remained unchanged. The Northwest Territories saw a decline in the rate from P1 to P2. Nova Scotia had the highest rate of uptake among uninsured children in P2 (717.8/1000), followed by the Northwest Territories (573.2/1000), Saskatchewan (528.3/1000), Nunavut (502.7/1000), and Manitoba (473.7/1000) (Figure 1c). Rates of child participation for the additional funding streams, AP1 and AP2 were also calculated (Table 2). Saskatchewan (9.6/1000 in AP1 and 15.2/1000 in AP2), Manitoba (7.7/1000 in AP1 and 13.9/1000 in AP2), Nunavut (8.0/1000 in AP1 and 13.1/1000 in AP2), and Nova Scotia (6.6/1000 in AP1 and 11.1/1000 in AP2) had the highest rates of applications for additional funding to cover the costs of their children’s dental care.

Figure 2 presents the distribution of children approved along with the total funds dispersed for the various periods according to age of the child. The greatest number of children with the Interim CDB in P1 and P2 were among those six years of age, while the highest amounts dispersed were to those five years of age and above (Figure 2a). The number of infants who received the Interim CDB increased from P1 to P2; 19.8% increase among those < 12 months of age (11,730 to 14,620) and 7.1% increase among one-year olds. There was also a 5.4% and 4.9% increase among two- and three-year-olds, respectively. Similarly, there was an increase in the amount of funding dispersed from P1 to P2 among infants; 20.1% increase among those < 12 months of age ($7,386M to $9.245M) and 8.6% increase among one-year olds ($11.7M to $12.7M). There was also a 5.8% and 5.5% increase among two- and three-year-olds, respectively. Meanwhile, there was a 4.7% decrease in the number of 11-year-olds who received the Interim CDB in P2 than in P1 (28,230 to 26,910) and a 4.2% decrease in the amount of funding provided to 11-year-olds ($17.2M to $16.5M). Figure 2b presents the distribution of the number of children approved and the total funds dispersed during AP1 and AP2. Overall, there was a 59.4% increase in the total number of approved children, rising from 24,660 in AP1 to 39,300 in AP2. Similarly, the total funding dispersed increased by 62.9%, from $15.1M to $24.6M. This upward trend was generally consistent across most age groups. However, there was a 51.1% decrease in the number of 11-year-olds approved (5,420 in AP1 to 2,650 in AP2). Additionally, the total funding for this age group declined from $3.4M to $1.6M.

**Figure 2.**
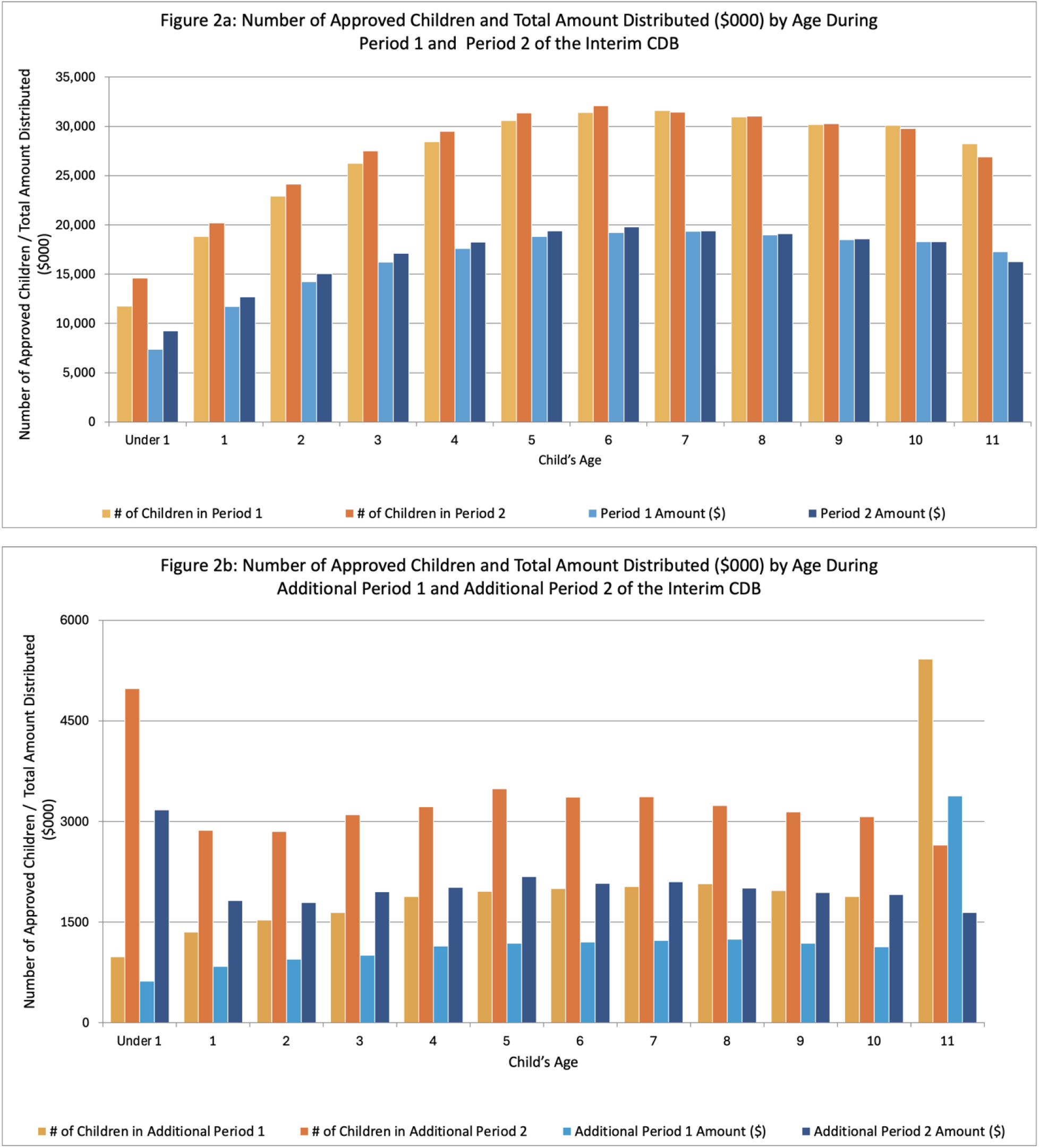
Total number of approved children and total amount distributed (in $000) during Period 1, Period 2, Additional Period 1, and Additional Period 2 of the Interim CDB

Table 3 reports the number of approved Interim CDB applications, unique applicants, children, and the total amount (in $000) by adjusted annual net family income during P1, P2, AP1, and AP2. In P1, 46.9% of approved children came from families with adjusted net incomes < $30,000. 44.5% were from families earning between $30,000 and $69,999, and only 8.9% were from families earning ≥ $70,000. During P2, the percentage of children from lower-income families (< $30,000) increased to 55.2%, while 37.6% were from families earning between $30,000 and $69,999, and 7.4% were from families earning ≥ $70,000. More than half of children (51.8%) approved in AP1 were from households earning < $30,000, which increased to 63.5% during AP2.

Information on the age grouping and place of residence of applicants appears in Table 4. The total number of applicants remained stable over the two periods, with 188,510 in P1 and 188,280 in P2 although P1 only consisted of 9 months while P2 was a 12-month span. Most approved applicants belonged to the 25-34 year old (36.5% P1 and 38.6% P2) and 35-44 year old (47.2% P1 and 46.1% P2) groups. This distribution was exhibited in all the provinces and territories. For example, in Ontario, 36.6% of parents in P1 were aged 25-34, increasing to 38.4% in P2, while those aged 35-44 accounted for 47.4% and 46.6%, respectively. In contrast, individuals < 25 and those aged ≥ 65 made up a minimal portion of applicants. For instance, in Newfoundland and Labrador, those under 25 comprised 6.4% in P1 and 5.1% in P2, while those aged 65 and older represented less than 1% across most provinces.

The actual number of applications, number of unique applicants, number of children receiving the benefit and total amount dispersed for the Interim CDB during the entirety of P2 were compared to projected values based on the first 9 months of the program’s second year.^8^ Overall, the actual number of applications and unique applicants at the end of P2 fell short of projections (Table 5), with 26,910 fewer applications and 26,747 fewer unique applicants. Similarly, 48,133 fewer children received the Interim CDB and $30.4M less dispersed than projected in P2.

## 4 Discussion

The Interim CDB was a temporary program providing financial relief to cover dental expenses for children < 12 years of age while the federal government planned the new CDCP. Our analyses of aggregate data for the entirety of the program reveals several key insights into the program’s uptake and impact on accessibility for dental care. As reported in our publications on the first 9 and 18 months of the program^8, 9^, significant regional and socio-economic variability exist, highlighting successes and areas for further improvement.

Ontario, Quebec, Alberta, and British Columbia had the highest number of approved applications, reflecting their population sizes and potentially greater socio-economic challenges. To control for the size differences in provinces and territories, rates of child participation were calculated to facilitate comparisons between provinces and territories. Those provinces and territories with the highest unadjusted uptake rates during the entirety of the Interim CDB included Manitoba, Ontario, Nova Scotia, and Saskatchewan, which confirms what we reported after reviewing data after the first 9 and 18 months of the program.^8, 9^ The higher participation in Manitoba and Saskatchewan may be due to reported poorer access to dental services as compared to other provinces. ^8, 12^ These two provinces were identified as having more notable access to oral health care challenges according to national surveys.^13^

After adjusting for the lack of private or public insurance, two provinces and one territory had the highest rates in both P1 and P2, namely Nova Scotia, Northwest Territories, and Saskatchewan. Manitoba also had higher rates of uptake than the other remaining provinces and territories. Rates in Nova Scotia, the Northwest Territories, and Saskatchewan were 2.0 times, 1.8 times, and 1.5 times the Canadian average, respectively. Uptake rates also increased markedly in Nunavut from P1 (395.0/1000) to P2 (502.7/1000) suggesting that targeted outreach efforts may have contributed to enhanced access in more remote locations. The high rates from Nova Scotia were surprising considering that the province has a legislated children’s dental public health program.^14^ However, the high rates of uptake from the Northwest Territories, Saskatchewan, and Manitoba may be attributed to the absence of provincial or territorial dental programs for children.^14^ According to data provided by Statistics Canada for this study, the estimated proportion of children 0-17 years of age without private and/or public dental insurance is 10.9% in Nova Scotia, 9.7% in the Northwest Territories, 14.6% in Saskatchewan, and 17.8% in Manitoba. Quebec and Yukon had the lowest uptake rates, 3.4 and 4.7 times lower than Nova Scotia’s and 1.7 and 2.4 times lower than the Canadian average after adjusting for uninsured children. Both regions offer public dental health programs for children.^14^ Data provided by Statistics Canada for this study reveal provincial and territorial differences in private and public insurance coverage among children aged 0-17. For instance, only 73.6% of children in Quebec had private and/or public dental insurance while over 85% or more of the pediatric populations in Saskatchewan, Yukon, Nova Scotia, Northwest Territories, and Nunavut had private or public dental insurance. This proportion of children with private and/or public dental coverage was also below 80% in New Brunswick, British Columbia, and Ontario. Quebec’s lower uptake rates were surprising and may signify potential challenges in awareness or accessibility along with the potential need for improved dialogue and cooperation between the federal and provincial governments in the oral health realm.

Income levels significantly influenced the uptake of the Interim CDB. Overall, as family income increased, the number of applicants fell sharply revealing that the Interim CDB primarily supported lower-income families. Families earning less than <$30,000 represented the majority of approved applicants across all periods, with participation increasing over time. Conversely, participation among high-income families ($70,000+) was consistently lower, indicating that the Interim CDB primarily supported those with the greatest need. However, lower uptake among higher-income families may also point to a perceived lack of necessity for the benefit among these groups. It raises an important question of whether families in this income bracket require government support for dental care.

Overall, the number of approved applications remained stable between P1 and P2, indicating steady demand for the Interim CDB. Regional shifts were evident, with Ontario and Quebec reporting the highest participation but a slight decline in Quebec from 18.2% in P1 to 17.4% in P2. The steady distribution amounts between periods highlight consistent support provided to families across Canada. One unique finding was the spike in AP2 for children under 1 year, as the number of children under one who received the AP2 payment for the Interim CDB is slightly higher than other age groups. Infant dental care is often more preventative than curative. It does not result in inordinately higher expenses - the most likely explanation is that children eligible for the benefit could only receive a maximum of two payments. Many children < 1 year would not have qualified for either the P1 or AP1 payments, increasing the likelihood of them applying for the AP2 additional payment to receive their maximum of two payments. For older children who had the option of receiving a P1 payment, there would be a reduced likelihood of applying for a P2 additional payment, as they were more likely to have exhausted the maximum number of payments before getting to AP2. Legislatively, children under 1 were eligible for the P2 additional if they met the other criteria. It appears that many young children in Canada may have benefited from the Interim CDB. The encouraging uptake of the intervention among parents with infants indicates early progress in improving oral health outcomes and may represent an important step toward reducing the burden of early childhood caries in Canada.

The discrepancies between projected and actual numbers of children and funds distributed in the second year of the Interim CDB underline significant policy implications. Actual applications, unique applicants, children receiving benefits, and total funds dispersed during P2 fell short of projections based on data from the first 18 months of the program.^8^ This is most likely because the new CDCP was being promoted for children under 18 years during the final quarter of the Interim CDB, leading parents to possibly wait to apply for CDCP instead. The fact that the actual figures for AP2 exceeded projections suggests a higher-than-expected demand for the benefit. This emphasizes the need for adaptive funding policies that effectively respond to fluctuating needs. Lower participation rates among higher-income families suggests a lack of awareness. Targeted campaigns could help ensure that all eligible families are informed about and can access the available support. Higher-income families are often more likely to possess employer-based dental benefits, resulting in eligibility disqualifications. The program might have been less relevant for wealthier households with needs beyond the plan, such as orthodontic services.

Due to the nature of data available on the Interim CDB, this study’s limitations include a lack of specific insights on overarching trends and broad utilization due to high aggregation and low dataset granularity. Specifically, within-province inequities may stem from urban/rural differences in dental networks, Interim CDB program enrolment, and accessibility for remote populations. Since the Interim CDB was a benefit paid to families, no data exist on dental treatments provided to children, their costs, and reimbursement details, which limits a deeper understanding of the benefit’s impact. Participation rates were calculated from the under-12 population using the 2021 census. Migration and differing fertility rates over the subsequent three years may have influenced the analysis.

Insurance coverage rates were calculated specifically for children aged 0–11 based on data originally collected for children aged 0–17. Thus, assuming a uniform distribution of insurance coverage across these age groups may have introduced inaccuracies. Actual coverage rates for younger children may have differed from these estimates, potentially affecting the precision of the analyses.

## 5 Conclusion

The Interim Canada Dental Benefit (CDB) has increased dental care access for low-income families with children under 12, particularly in high-need areas. However, uptake disparities persist across regions, income levels, and age groups, necessitating further interventions. Enhancing accessibility and tailoring strategies to regional needs will be vital for equitable distribution. Building on this, the Canadian Dental Care Plan (CDCP) aims to expand coverage for up to nine million uninsured Canadians, easing financial barriers and reducing disparities for vulnerable groups. ^14^ These initiatives mark a significant shift in Canada’s dental health policy, targeting underserved communities with critical funding. Ensuring resources meet increased demand, particularly in underserved areas and during benefit periods, is essential for the program’s effectiveness and reach.

## 7 Author Contributions

RJS and AM conceptualized the work. RJS led data curation and project administration. RJS, VCJ, CY, OOO, VHKL, SG, and AM contributed to data analysis and interpretation. All authors contributed to the draft of the manuscript and critically reviewed it. All authors contributed to the article and approved the submitted version.

## 9 Acknowledgments

Dr. Robert J Schroth holds a Canadian Institutes of Health Research Applied Public Health Chair in “Public Health Approaches to Improve Access to Oral Health Care and Oral Health Status for Young Children in Canada”.

## 11 Data Availability Statement

The datasets analyzed for this study can be found in the Government of Canada Open Data Portal: https://open.canada.ca/data/en/dataset/69035265-2714-4ffa-af3f-fa850209b616 and https://www150.statcan.gc.ca/t1/tbl1/en/cv.action?pid=1310092001.

